# Anticoagulation Usage and Thrombolytic Therapy in Subjects with Atrial Fibrillation Associated Ischaemic Stroke

**DOI:** 10.1101/2024.07.28.24311145

**Authors:** J Harbison, J McCormack, O Brych, R Collins, N O’Connell, P Kelly, T Cassidy

**Affiliations:** Irish National Audit of Stroke, National Office of Clinical Audit. St Stephen’s Green, Dublin 2, Ireland; Dept of Medical Gerontology, School of Medicine, Trinity College Dublin, Dublin 2, Ireland; National Coagulation Centre, St James’s Hospital, Dublin 8, Ireland; Dept of Neurology, University College Dublin. Mater Hospital Institute of Neurology, Dublin 1, Ireland; Acute Stroke Service, St Vincent’s Hospital, Elm Park Dublin 4, Ireland

**Author notes:** Corresponding Author. Prof Joseph Harbison., Department of Medical Gerontology, Trinity College Dublin, Mercer’s Institute for Successful Ageing, St James’s Hospital, Dublin 8.

## Abstract

Atrial Fibrillation (AF) has been Identified as the single largest cause of ischaemic stroke in the Irish population. Previous research showed that widespread adoption of Direct Oral Anticoagulant (DOAC) prophylaxis had not been associated with a reduction in AF associated stroke prevalence. The Irish National Audit of Stroke (INAS) undertook a more detailed study to determine the characteristics of anticoagulation practice in AF associated stroke particularly adherence to prescribing guidelines and effect on thrombolysis rate.

**Methods:** Data from INAS were analysed for the period 2017-2022 inclusive as part of a cross-sectional, cohort study. An enhanced dataset with additional questions about adherence with prescription and International Normalized Ratio (INR) control was collected for 2022 was also considered separately.

**Results:** Complete AF Data were available on 22485 of 26829 incidents admitted over this period. Of these, 19260 (85.6%) were ischaemic strokes, mean age was 71.8 and 57.1% were male. In 5321 of these cases, the AF was identified and in 2835 (53.3%) recognized before the stroke and 2281 (80.4%) had been prescribed anticoagulation. The population with previously unknown AF were significantly younger on average than those on anticoagulation (76.8 years vs. 79.1 years (p<0.0001, t-test)), Group C; 78.8 years (p<0.0001), they were also much more likely to have received thrombolysis (17.3 % vs. 4.0% (Chi Sq, p<0.0001)).

There were 4999 stroke in 2022, 4272 (85.4%) were ischemic and 1270 (29.7%) of these were AF associated. Of the 660 total strokes, 597 (90.5%) anticoagulated at presentation were receiving DOACs, of which 557 were ischaemic. Forty-eight (9.5%) had their anticoagulation paused and 40 admitted to poor compliance (7.9%).

**Conclusion:** Nearly half of people with AF detected after stroke was previously unknown. Those with known AF were mainly appropriately treated with DOACs and constitute breakthrough strokes. Subjects receiving DOACs were much less likely to receive thrombolytic therapy even than those taking Warfarin.

## Introduction

Atrial Fibrillation (AF) is a major cause of Ischaemic Stroke (1) and has previously been identified as the single biggest cause of ischaemic stroke in the Irish population (2). In 2022 research by the Irish National Audit of Stroke (INAS) and The National Centre for Pharmacoeconomics (NCPE) showed that whilst there had been a substantial increase in people receiving anticoagulation therapy since the advent of Direct Oral Anticoagulant (DOAC) therapy, this did not seem to be matched by a reduction in the proportion of AF associated strokes (3). There were a number of potential causes for this including issues with adherence to anticoagulant prescription, incorrect prescription of the medications or simply a lag in response to. In light of this, INAS undertook a further review of the data and requested centres to collect an additional data on AF and anticoagulation for the calendar year 2022 to examine more closely prescribing practices adherence with therapy and to determine if they were less optimal that in other populations (4). In addition, INAS had noted a small reduction in the proportion of patients receiving thrombolysis in the preceding few years and it was hypothesised that an increase in the use of DOAC therapy, an absolute contraindication to thrombolysis, may have had an influence on proportion of people thrombolysed (5).

## Methods

Data were obtained from the Irish National Audit of Stroke (INAS) as part of a cross-sectional, cohort study. This is a dataset routinely collected by clinical stroke teams in all hospitals in Ireland admitting at least 25 stroke patients per annum (6). The data is collected, primary for quality improvement purposes, to a national online database where it is merged with routinely collected demographic, process and outcome data from the Hospital In-Patient Enquiry (HIPE). Only data from hospitals where there is at least 80% agreement between INAS and HIPE datasets are included. Data on subarachnoid haemorrhage is not routinely collected or reported by the audit as admission pathways differ than for ischaemic stroke and Primary Intracerebral Haemorrhage.

The Audit was established in 2013 and data is stored in an anonymised form. Compared to other registers and audits, INAS has an emphasis on Atrial Fibrillation (AF) and Anticoagulation status due to the high incidence of atrial fibrillation associated strokes previously identified in Ireland. Therefore, in 2022 an additional snap-shot audit was performed, with supplementary data items added to the dataset, in response to a study showing little evidence of improved stroke outcome despite substantial investment in DOAC therapy nationally.

Data was analysed for the 6-year period 2017-2022 inclusive to determine the prevalence of AF in the population of admitted strokes and particularly in those with known AF prior to their stroke. Data was also analysed to evaluate the use of anticoagulant therapy and to determine if populations receiving pre-stroke anticoagulation differed and whether anti-coagulation status was associated with differences in outcome including length of stay, disability as indicated by the modified Rankin Score, and with mortality.

Data were further analysed from the 2022 snapshot audit to examine the reported level of adherence with anticoagulation therapy of patients prior to stroke and to what extent AF associated, ischaemic strokes occurring while on anticoagulation were due to inadequate or ineffective therapy.

Data were analysed using proprietary statistical software and using Microsoft Excel. Quantitive data were analysed using student’s t-tests and proportional data using chi square statistics. Although all data were anonymised prior to analysis, ethical approval was obtained for the study from the joint St James’s Hospital /Tallaght University Hospital institutional ethics committee (ref 3989) to comply with institutional practice for INAS where data is used outside standard outcome reporting.

## Results

From 2017 to 2022, 26829 individual stroke events were recorded on the audit database of which 23051 (85.9%) were ischaemic. Full AF data were available on 22484 (83.8%) (Figure 1) of which 19260 (85.7%) were ischaemic. Mean age of subjects with full AF data was 71.8 years (s.d. 13.8) at the time of stroke and 12857 (57.1%) were male. Over one quarter (n=5920, 26.3%) of the group with AF data had a confirmed diagnosis of AF, and of these 5321 (89.9%) were ischaemic strokes. In 2835 (53.3%) of these cases, the AF was diagnosed before the stroke and of these 2281 (80.4%) had been prescribed anticoagulation prior to stroke.

**Figure 1.**
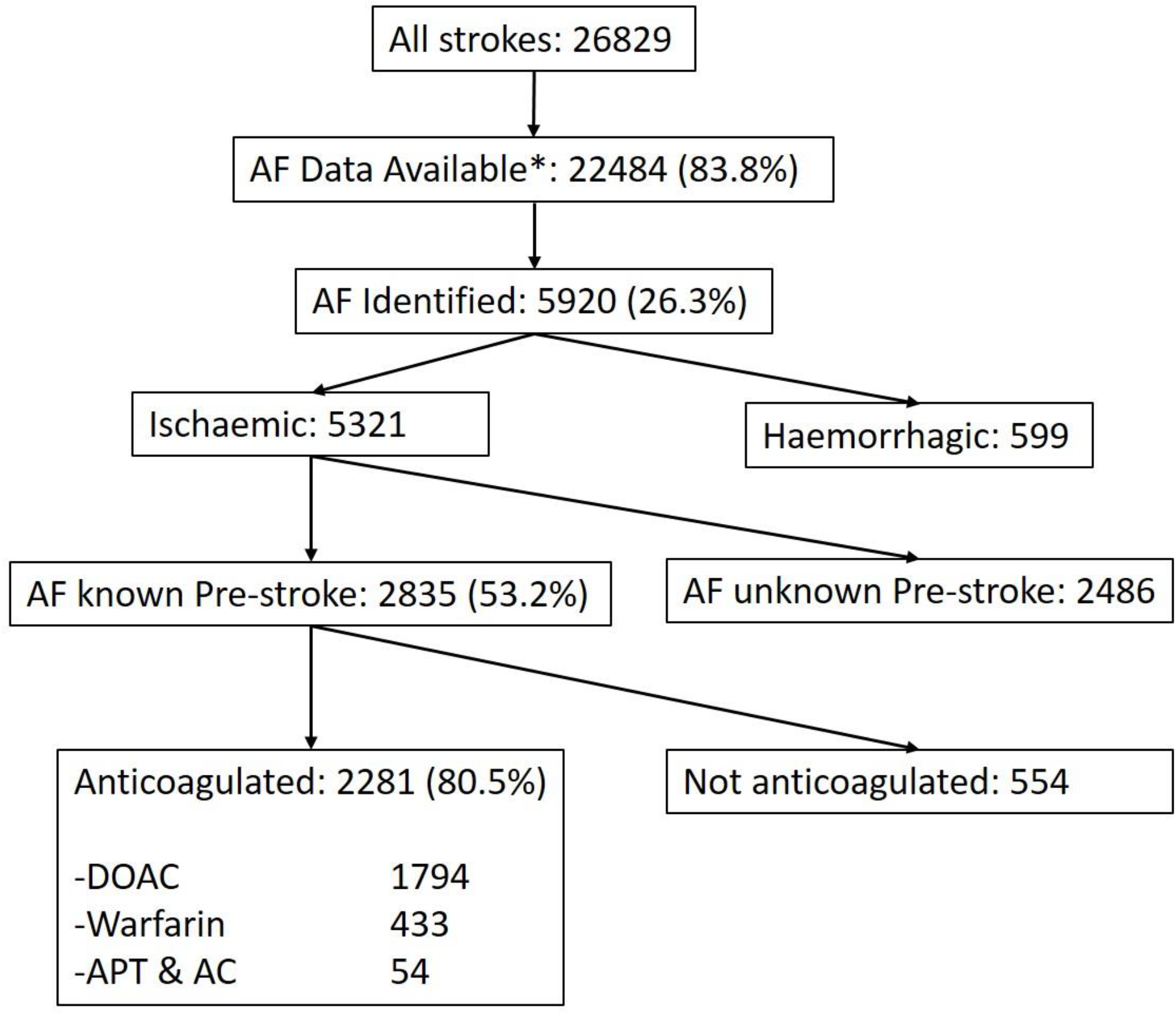
Flow chart of admissions with Stroke and detected Atrial Fibrillation 2017-2022 from Irish National Audit of Stroke. *Patents with no data missing or not listed as still under investigation.

The data on subjects with ischaemic stroke and AF were divided into those with unknown AF at stroke onset (Group A: n=2486), those with known AF at onset who had been anticoagulated (Group B: n=2281) and those with known AF at onset who were not anticoagulated (Group C: n=554) for analysis (Table 1). Comparisons for continuous data were made using Student’s t tests and for proportional data using Pearson’s Chi Square statistics (Table 1).

**Table 1:** Characteristics and Outcomes in Anticoagulation Subgroups.

|  | A:<br>Previously<br>Unknown<br>Atrial<br>Fibrillation | B:<br>Known Atrial<br>Fibrillation<br>Anticoagulated | P<br>A vs. B | C:<br>Known Atrial<br>Fibrillation<br>Not<br>Anticoagulated | P<br>A vs C | P<br>B vs. C |
| --- | --- | --- | --- | --- | --- | --- |
| N = 5321 | 2486<br>(46.7%) | 2281<br>(42.9%) |  | 554<br>(10.4%) |  |  |
| Mean age,<br>years<br>(SD) | 76.8<br>(11) | 79.1<br>(9) | <0.0001 | 78.8<br>(11) | <0.0001 | 0.51 |
| Male % | 54.1% | 58.4% | 0.003 | 55.4% | 0.58 | 0.2 |
| Median<br>Length of<br>stay, days<br>(range) | 11<br>(1-552) |  | 10<br>(1-534) |  | 10<br>(1-407) |  |
| Length of<br>stay <7days<br>(%) | 29.7% | 31.8% | 0.11 | 33.0% | 0.12 | 0.58 |
| Thrombolysis<br>rate<br>(%) | 17.3% | 4.0% | <0.0001 | 14.1% | 0.07 | <0.0001 |
| Pre-Stroke<br>mRS data<br>available n<br>(%) | 2160<br>(86.9%) | 2073 (90.9%) |  | 512 (92.4%) |  |  |
| Pre-Stroke<br>mRS<br>0-1 | 77.3% | 63.9% | <0.0001 | 56.6% | <0.0001 | 0.002 |
| Discharge<br>mRS n (%) | 2139<br>(86.0%) | 2058<br>(90.2%) |  | 502<br>(90.6%) |  |  |
| Discharge<br>mRS<br>0-1 (%) | 34.5% | 28.6% | <0.0001 | 26.2% | 0.0004 | 0.3 |
| Discharge<br>mRS<br>6 (%) | 9.7% | 13.2% | <0.0003 | 19.7% | <0.0001 | 0.0002 |

mRS: Modified Rankin Score

The population with previously unknown AF were significantly younger on average (Group A: 76.8 years, Group B 79.1 years (p<0.0001, t-test), Group C; 78.8 years (p<0.0001). This appears to be due to a higher proportion of under 70s in Group A (<70 years: Group A 23.3%, Group B + C 13.5% (Chi Sq 86.2, p<0.0001). While a majority in all groups were male, there was a significantly higher proportion of men in the group that was anticoagulated (Group B vs. Group A and Group C) 58.4% vs. 54.3% (chi sq 8.5, p=0.004).

In outcomes, there was no significant difference in length of stay between groups. Data was missing on admission and discharge modified Rankin score in between 8% and 14% of records., patients admitted with stroke and previously unrecognised AF (Group A) had significantly better outcomes in terms of independent recovery and mortality (Table 1).

Thrombolysis practice differed greatly between groups. Where 17.3 % of Group A were thrombolysed and 14.1 % of group C only 4.0% of the anticoagulated Group B were thrombolysed (p<0.0001). This data was further analysed to compare those prescribed only DOACs (n=1794) and these receiving Vitamin K antagonists (warfarin) (n=433). Of the DOAC group only 52 received thrombolysis thrombolysed (2.9%) compared with 37 (8.5%) of the VKA Group (Chi Sq 29.0 p<0.0001).

There were 4999 stroke episodes recorded in the 2022 group with supplementary data (Figure 2), 4272 (85.5%) were ischaemic. Proportionally, 29.7% (n=1270) of the 4272 ischemic strokes, and 20.6% (n=150) of the 727 haemorrhagic strokes were associated with AF (Chi Sq 4.23, p=0.04). AF was previously known in 785 of the ischaemic strokes (61.8%) and 133 of the haemorrhagic strokes (86.7%) (Chi Sq 42.3, p<0.0001). Of these, 557 of the ischaemic strokes (70.9%) and 103 of the haemorrhagic strokes (77.4%) with known AF were anticoagulated (Chi Sq 2.37, p=0.12).

**Figure 2:**
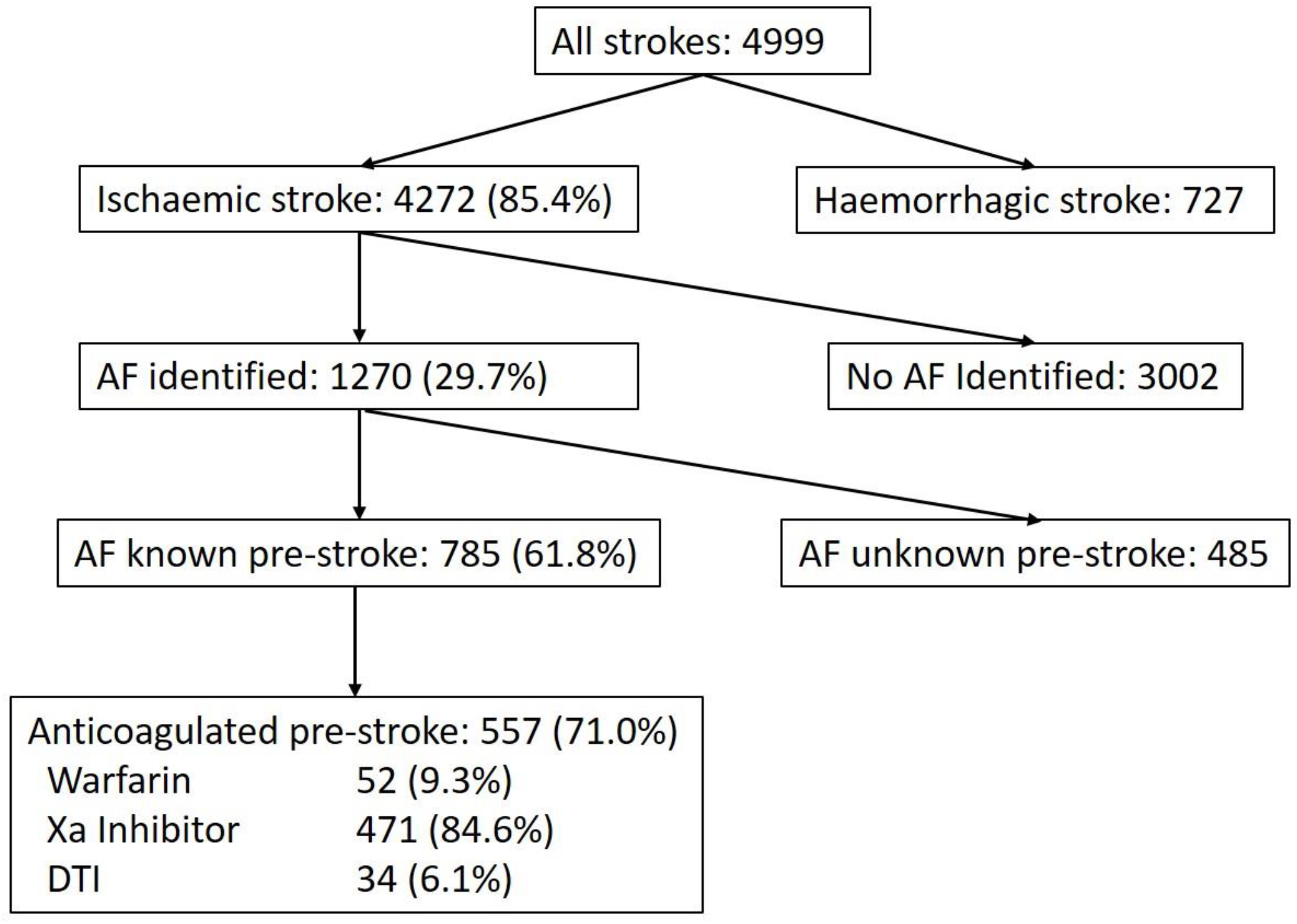
Admissions during enhanced data collection, January to December 2022. Xa Inhibitor: Factor Xa Inhibitor, Direct Oral Anticoagulant DTI: Direct Thrombin Inhibitor, Direct Oral Anticoagulant.

Of the 660 anticoagulated patients 597 (90.5%) were prescribed DOACs, and 473 (79.2%) of these subjects were confirmed as receiving the correct dose. Only 25 (39.7%) of the 63 patients receiving Warfarin had an INR in therapeutic range on admission. Of the 505 subjects with AF and Ischaemic stroke who had been prescribed DOACs, 48 (9.5%) had their anticoagulation paused at time of stroke, 19 (39.6%) of these because of a planned medical or dental procedure. Almost three quarters (n=376, 71.3%) reported always taking their prescribed medicine, and 40 admitted to poor compliance (7.9%). Adherence data was unavailable for 105 patients (20.8%).

## Discussion

In this national study, more than one quarter of subjects with ischaemic stroke were ultimately found to have AF. Of these, almost a half were unaware of the AF at time of stroke. Whilst the proportion of males were higher in all groups, the difference in prevalence of AF between males and females was significantly smaller in the group with previously unrecognised group. The group with previously unrecognised AF were also significantly younger on average.

More than two fifths of patients with known AF who had ischaemic strokes were anticoagulated at presentation however, this cohort had worse outcomes than those who were not anticoagulated. Our analysis of the 2022 data showed that four fifths of those prescribed DOACs were found to be on the correct dose whereas only two fifths of those on Warfarin were in therapeutic range. The majority of strokes in patients on DOAC therapy were in people taking the correct dose and with good self-reported adherence to prescription. Subjects who were anticoagulated, especially those prescribed DOACs, were far less likely to be thrombolysed. Indeed, evaluation of the 2022 data showed that those found to be receiving an appropriate dose of DOAC at presentation were nearly never thrombolysed.

This study has some limitations. Whilst the audit collects data from all hospitals with stroke units admitting stroke patients, there are patients who are not identified, including those admitted to smaller hospitals, those admitted to private institutions and those managed in community settings. There were also some years where hospitals failed to meet the 80% standard of agreement with HIPE data and they would also have been excluded however, the number of excluded cases was low in comparison with the National figures. The data is predominantly collected by Clinical Nurse Specialists in Stroke from patient records and reflects care and practice in hospitals data in some indices can be less complete than others. In this study this was particularly the case with modified Rankin Score on discharge where 14% of the patients were missing this data, frequently because the assessment was not made. It should be noted that data on modified Rankin score of 6 (death), is doubly collected in routing patient outcome data and is complete. We do not believe that it is likely there was a differential effect on collection of this data based on anticoagulation status though, and thus feel that the difference in outcome found is reliable. Previous comparison of data quality from continuous audit against chart based audit has found close agreement (7).

Our finding that the population with previously unrecognised AF were younger due to a higher proportion of under 70 year olds in the group is interesting because of the potential role of opportunistic AF screening (8). People over 70 years of age in Ireland are entitled to free General Practitioner visits in the community and a policy of opportunistic screening for Atrial Fibrillation for AF is recommended and has been widely practiced (9). It was formally introduced as part of a Chronic Disease Management programme in 2020 (10). This is not routinely available to under 70 year olds, the majority of whom must pay for GP visits.

A high proportion of patients (>80%) with known AF had been commenced on appropriate Anticoagulation therapy. Analysis of the 2022 data showed that the great majority were receiving DOAC therapy and of these, nearly 80% were taking the correct dose according to prescribing guidelines. This level of adherence to therapy is consistent with previous reports from other centres (11, 12, 13). This implies that the majority of ischaemic strokes on DOAC therapy were so-called ‘breakthrough’ strokes (14, 15), the reasons for which are varied. INAS offered services the facility of checking DOAC levels on patients through the National Coagulation Centre, in St James’s Hospital but it was rarely accessed. The proportion of the smaller number of individuals on Warfarin therapy who were in therapeutic range at the time of their presentation was in contrast <50%. Ireland has historically had a limited number of anticoagulation clinics, especially in non-urban areas (16) that the widespread abandonment of warfarin in favour of a DOAC may have resulted in reduced familiarity with and effectiveness of Warfarin monitoring. A remarkable finding of this study is the apparent effect that different anticoagulation strategies had on likelihood of thrombolysis. Those who had AF and were not anticoagulated had a thrombolysis rate of >15% but this dropped to just over 4% in anticoagulated patients. examination of the 2022 data showed that those admitted on Warfarin had nearly three times the thrombolysis rate than those on DOACs and subjects who were determined to be adherent to therapy and on the correct dose of anticoagulant were almost never thrombolysed at all. This may reflect the fact that guidelines will permit thrombolysis on Warfarin therapy with an INR of ≤1.8 but DOAC therapy typically remains a complete contraindication. Therapeutic monitoring of drug levels is possible for DOACs and, although offered in this study, it was rarely availed of and did not change treatment decisions it respect of thrombolysis. It is of note that a number of observational studies (17, 18, 19) have found that thrombolysis of patients on DOAC therapy is not associated with worsening outcome. No patient who was on DOAC therapy apparently received a reversal agent to permit thrombolysis although many would have been referred for thrombectomy through the National Thrombectomy Service centres (20). It is plausible that the increasing proportion of AF associated strokes presenting in patients on DOAC therapy may be influencing the overall rate of thrombolysis and if generalised internationally these may represent a substantial number of individuals not-receiving an effective acute stroke therapy and there may be a case for a randomized trial in this group.

In conclusion, the majority of subjects admitted with AF associated stroke had their AF recognised before the acute event and the majority of these were appropriately anticoagulated, highlighting the substantial issue of breakthrough strokes and their prevention. Patients who were treated with DOAC therapy at presentation were significantly less likely to receive thrombolysis therapy than those who were untreated or even those who were anticoagulated using warfarin. Consideration should be given to whether evidence supports a complete proscription of thrombolysis therapy in subjects presenting with breakthrough stroke on DOAC therapy (21).

## Data Availability

All data is derived from the Irish National Audit of Stroke. This is a publicly available data source.

